# Integrated NMR and MS analysis of plasma metabolome reveals major changes in inflammatory markers, one-carbon, lipid, and amino acid metabolism in severe and fatal COVID-19 subjects

**DOI:** 10.1101/2023.04.19.23288802

**Authors:** Marcos C. Gama-Almeida, Lívia Teixeira, Eugenio D. Hottz, Paula Ivens, Hygor Ribeiro, Gabriela D. A. Pinto, Rafael Garrett, Alexandre G. Torres, Talita I. A. Carneiro, Bianca de O Barbalho, Christian Ludwig, Claudio J. Struchiner, Iranaia Assunção-Miranda, Ana Paula C. Valente, Fernando A. Bozza, Patrícia T. Bozza, Gilson C. dos Santos, Tatiana El-Bacha

## Abstract

Brazil has the second highest COVID-19 death rate while Rio de Janeiro is among the states with the highest rate in the country. Although effective vaccines have been developed, it is anticipated that the ongoing COVID-19 pandemic will transition into an endemic state. Under this scenario, it is worrisome that the underlying molecular mechanisms associated with the disease clinical evolution from mild to severe, as well as the mechanisms leading to long COVID are not yet fully understood. In this study, ^1^H Nuclear Magnetic Resonance spectroscopy and Liquid Chromatography-Mass spectrometry-based metabolomics were used to identify potential pathways and metabolites involved in COVID-19 pathophysiology and disease outcome. We prospectively enrolled 35 severe RT-PCR confirmed COVID-19 cases within 72 hours from intensive care unit admission, between April and July 2020 from two reference centers in Rio de Janeiro, and 12 samples from non-infected control subjects. Of the 35 samples from COVID-19 patients, 18 were from survivors and 17 from non-survivors. We observed that patients with severe COVID-19 had their plasma metabolome significantly changed if compared to control subjects. We observed lower levels of glycerophosphocholine and other choline-related metabolites, serine, glycine, and betaine, indicating a dysregulation in methyl donors and one-carbon metabolism. Importantly, non-survivors had higher levels of creatine/creatinine, 4-hydroxyproline, gluconic acid and *N*-acetylserine compared to survivors and controls, reflecting uncontrolled inflammation, liver and kidney dysfunction, and insulin resistance in these patients. Lipoprotein dynamics and amino acid metabolism were also altered in severe COVID-19 subjects. Several changes were greater in women, thus patient’s sex should be considered in pandemic surveillance to achieve better disease stratification and improve outcomes. The incidence of severe outcome after hospital discharge is very high in Brazil, thus these metabolic alterations may be used to monitor patients’ organs and tissues and to understand the pathophysiology of long-post COVID-19.

## Introduction

The existing vaccines for SARS-CoV-2 infection resulted in a significant reduction in the number of severe cases of the disease. However, it is anticipated that the ongoing coronavirus disease 2019 (COVID-19) pandemic will transition into an endemic state [1]. By the time this article is being written, COVID-19 has exceeded 762 million cases with almost 6.9 million deaths worldwide [2]. Brazil has recorded more than 700,000 deaths, making it the country with the second highest number of deaths worldwide [2]. Rio de Janeiro, where this study took place, had 2.79 million confirmed cases with approximately 77,000 deaths. The state is among the ones with the highest mortality rate [3].

The replication of SARS-CoV-2 triggers a systemic immune response that leads to tissue damage and reprogramming of whole-body metabolism [4,5]. Additionally, around 20 % of infected subjects may experience long-term symptoms after recovery from the initial illness [6], a condition that is associated with neurological, gastrointestinal, pulmonary, and cardiovascular alterations and which can be highly debilitating [7]. Even patients who present mild symptoms in the acute phase of the disease may later develop post-COVID symptoms [8].

These observations reveal the complexity of COVID-19 pathophysiology and its profound impact on different tissues and cells. However, the underlying molecular mechanisms associated with the disease clinical evolution from mild to severe, as well as the mechanisms leading to long COVID-19 symptoms are not yet known. The complex multisystemic nature of SARS-CoV-2 infection calls for a system-level approach to help us better understand the underlying molecular mechanisms associated with the condition.

Metabolomics is the central omics in information translation [9] by providing the metabolic signature of organs and biological fluids in different conditions and may help elucidate the metabolic pathways associated with SARS-CoV-2 infectious process. We and others have shown that metabolomics was essential in the development of new approaches that improved the understanding and treatment of emerging viral diseases such as Dengue [10,11], Chikungunya [10], SARS [12], and Zika [13,14]. Alterations in host energy, amino acid and lipid metabolism are frequently observed in viral infections, as the virus disturbs and exploits host metabolic pathways for its own benefit [15,16]. These metabolic alterations have a critical role in disease outcome and in modulating the host immune response.

The modulation of host lipid metabolism is a feature shared by coronaviruses and is essential for viral RNA replication [17], as it enables the viral envelope membrane, as well as double-membrane vesicles and lipid compartments to be synthetized. Indeed, our group has shown that lipid droplets accumulate in monocytes isolated from COVID-19 subjects and serve as an assembly platform for SARS-CoV-2 particles [18]. The orchestrate of lipid flow within different cell compartments by the SARS-CoV-2 non-structural protein 6 (NSP6) ensures the proper organization of double-membrane vesicles, as well as their effective communication with lipid droplets [19]. All these events are essential for SARS-CoV-2 replication.

One of the first studies to use a multi-omics approach to gain insight into the pathophysiology of COVID-19 was done by Shen *et al*. with a small cohort of patients. In the study, proteomics and metabolomics approaches revealed that mild and severe COVID-19 patients presented metabolic and immune dysregulation [20]. More than 100 lipid species, including glycerophospholipids, fatty acids, and lipoproteins, and several amino acids, were downregulated in sera from subjects infected with SARS-CoV-2, if compared to controls [20]. Alterations in lipoproteins using NMR-metabolomics were also confirmed in larger cohorts [21] and single-cell metabolomics of monocytes reinforced the idea that modulation of intermediary metabolism, in particular organic acids, plays crucial roles in COVID-19 severity [22]. Regarding the modulation of amino acid metabolism, several studies have reported that patients have low levels of plasma tryptophan, being now considered a marker for the extent of inflammation and COVID-19 severity [20, 23-25]. Additionally, it has been reported that COVID-19 in-patients show dysregulation in the metabolism of methyl donors, including higher levels of S-adenosyl-homocysteine and lower levels of homocysteine, regardless of IL-6 levels [26]. Indeed, SARS-CoV-2 genome replication seems to depend on folate and methionine cycles modulation [27]. Therefore, SARS-CoV-2 infection is thought to affect various aspects of host metabolism and the extent of these changes is believed to be linked to the severity of the disease. On the other hand, the specific metabolic differences that may distinguish the severe cases of COVID-19 from those that are fatal have not yet been fully addressed.

In this study, we used ^1^H Nuclear Magnetic Resonance (NMR) spectroscopy and Liquid Chromatography-High-Resolution-Mass spectrometry (LC-HRMS)-based metabolomics to perform a thorough investigation of the metabolic disturbances induced by SARS-CoV-2 infection in a well characterized prospective cohort of subjects with severe COVID-19, including survivors and non-survivors, and healthy subjects. Patients’ samples were collected in Rio de Janeiro, Brazil, between April and July 2020. We were particularly interested in investigating metabolites associated with one-carbon metabolism and with lipid and amino acid metabolism.

The incidence of severe outcome after hospital discharge is very high in Brazil, thus these metabolic alterations may be used to monitor patients’ organs and tissues and to understand the pathophysiology of long-post COVID-19.

## Materials and Methods

### Study design and Participants

We prospectively enrolled severe RT-PCR confirmed cases within 72 hours from intensive care unit (ICU) admission in two reference centers at the Instituto Estadual do Cérebro Paulo Niemeyer and Hospital Copa Star, Rio de Janeiro, Brazil between April and July 2020. Adults (≥18 years of age) with severe COVID-19 (n=35) were enrolled in this study. Patients’ clinical status was confirmed by presence of chest infiltrates on computed tomography scan and by the need of respiratory support with either non-invasive oxygen supplementation or mechanical ventilation. The complete clinical information was collected prospectively using a standardized form: International Severe Acute Respiratory and Emerging Infection Consortium (ISARIC)/World Health Organization (WHO) Clinical Characterization Protocol for Severe Emerging Infections (CCP-BR). Upon admission, clinical and laboratory data were recorded for all severe patients included in the study. The primary outcome analyzed was 28-day mortality, and patients were classified as survivors (n=18) or non-survivors (n=17).

All ICU-admitted patients received the usual supportive care for severe COVID-19. Patients with acute respiratory distress syndrome (ARDS) were managed with neuromuscular blockade and a protective ventilation strategy that included low tidal volume (6 mL/kg predicted body weight) and limited driving pressure (<16 cmH_2_O) as well as optimal positive end-expiratory pressure calculated based on the best lung compliance and PaO_2_/fraction of inspired oxygen (FiO_2_) ratio. Antithrombotic prophylaxis was performed with 40 to 60 mg of enoxaparin per day. Patients did not receive antivirals, steroids, or other anti-inflammatory or antiplatelet drugs.

Peripheral blood was also collected from SARS-CoV-2 negative participants (control group; n=12) confirmed by RT-PCR of swabs on the day of blood sampling. The control group included subjects of matching age and sex distribution compared to infected subjects. These participants were not under anti-inflammatory or antiplatelet drugs for at least 2 weeks prior the study.

The National Review Board of Brazil approved the study protocol (Comissão Nacional de Ética em Pesquisa [CONEP] 30650420.4.1001.0008), and informed consent was obtained from all subjects or their caregivers.

### Chemicals and solvents

All solvents used were of HPLC analytical grade. Acetonitrile and methanol were obtained from TEDIA® (Fairfield, USA) and isopropanol from Sigma Aldrich (São Paulo, Brazil). Water was purified in the Milli-Q device, Millipore Purification System (Billerica, MA, USA). Mobile phase additives formic acid and ammonium hydroxide were purchased from TEDIA® and ammonium acetate was obtained from J. T. Baker® (Brazil). Isotopically labelled internal standards U^13^C D-glucose and U^13^C L-glutamine and deuterium oxide were purchased from Cambridge Isotope Laboratories, Inc. (MA, USA). All other standards were obtained from Sigma Aldrich.

### Sample processing

Blood samples were drawn into acid-citrate-dextrose and centrifuged (200 × *g*, 20 minutes, room temperature). Plasma was collected and stored at −80°C until analysis. Citrate-dextrose buffer was chosen to preserve platelets. This choice of buffer prevented us from comparing citrate and sugars among groups and limited the identification of metabolites that present chemical shifts in the ^1^H NMR spectrum, which is in the proximity of citrate.

### Nuclear Magnetic Resonance-based metabolomics

#### Sample preparation

Frozen plasma samples were quickly thawed and diluted 3-fold in sodium phosphate buffer and deuterium oxide (final concentration 50 mM phosphate buffer and 10 % deuterium oxide, pH 7.4). A total of 600 μL of diluted samples were transferred to a 5 mm NMR tube.

#### NMR acquisition and spectra pre-processing and metabolite assignment

NMR spectra were acquired on a Bruker Advance III 500.13 MHZ at 300 K, coupled with a cooled automatic sample case at 280 K. 1D-^1^H NMR spectra were acquired using excitation sculpting to suppress the solvent signal [28] as well as a CPMG (Carr-Purcell-Meiboom-Gill) T2 filter [29] with 32 loop counters and delay of 0.001 s. 32768 complex data points were acquired per transient with a total of 1024 transients. The spectral width was set to 19.99 ppm, resulting in an acquisition time of 3.27 s per FID. The relaxation delay was set to 1.74 s.

Spectra data were pre-processed in the MetaboLab [30] software. Prior to Fourier transform, the FIDs were apodised using an exponential window-function with 0.3 Hz line-broadening and then zero-filled to 65536 data points. After Fourier transform, each spectrum was manually phase corrected, followed by a spline-baseline correction. Finally, all spectra were referenced to the signal of the ^1^H linked to the anomeric carbon of glucose. Baseline noise and regions corresponding to water and citrate signals were deleted. Spectra data were binned with 0.005 ppm interval and transformed by the generalized logarithm function [31]. The resulting table presented 81,498 data points, corresponding to metabolites’ intensities.

Following 2D spectra, HSQC ^1^H-^13^C and TOCSY ^1^H-^1^H, acquisition, data was uploaded on the COLMAR [32,33] for the assignments. We also overlaid spectra from the BMRB [34] and HMDB 4.0 [35] databank. The software ICON NMR (Bruker) was used for automatic acquisition.

### Mass spectrometry-based metabolomics

#### Standards

A stock of internal standard (IS) solution was prepared with a final concentration of 0.15 mg mL^−1^ for U-^13^C D-glucose and 0.13 mg mL^−1^ for U-^13^C L-glutamine in acetonitrile/isopropanol/water (3:3:2, % v/v/v).

Stock solutions of targeted analytes (Supplementary Tables 1 and 2) were prepared at 1.0 mg mL^−1^ in methanol or in different proportions of acetonitrile/water. A standard working solution was prepared by mixing appropriate volumes of each stock solution to reach the final concentration of 2.0 – 50.0 µg mL^−1^ in acetonitrile/water (1:1, %/v).

**Table 1:** Demographics and clinical characteristics of control, COVID-19 survivors and non-survivors.

|  | **control**<br>**(n=12)** | **survivors**<br>**(n=18)** | **non-survivors**<br>**(n=17)** | ***P* value** |
| --- | --- | --- | --- | --- |
| Age, years | 50 (36–59) | 56 (39–63) | 58 (51–73) | 0.344 |
| Sex, male; *n* (%) | 5 (41 %) | 7 (38 %) | 10 (58 %) | 0.727 |
| Respiratory support; *n* (%) |  |  |  |  |
| Noninvasive O<sub>2</sub> supplementation | 0 (0 %) | 8 (44 %) | 0 (0 %) | **0.003** |
| Mechanical ventilation | 0 (0 %) | 10 (66 %) | 17 (100 %) |  |
| SAPS II | n.a. | 55 (37–64) | 68 (59 – 78.5) | **0.001** |
| PaO<sub>2</sub>/FiO<sub>2</sub> ratio | n.a. | 196 (154–429) | 139 (177–178) | 0.099 |
| Time from symptom onset to blood sample (days) | n.a. | 10 (7–14) | 10 (3– 14) | 0.975 |
| **Comorbidities; *n* (%)** |  |  |  |  |
| Obesity | 1 (8.3 %) | 5 (27.7 %) | 2 (11.7 %) | 0.650 |
| Hypertension | 2 (16%) | 4 (22%) | 5 (29 %) | 0.855 |
| Diabetes | 0 (0 %) | 6 (33 %) | 6 (35 %) | 0.990 |
| Cancer | 0 (0 %) | 2 (11 %) | 2 (11 %) | 0.990 |
| Heart disease<sup>1</sup> | 0 (0 %) | 2 (11 %) | 2 (11 %) | 0.990 |
| **Laboratory findings at admission** |  |  |  |  |
| Leukocytes, x 1000/μL | n.a. | 12.4 (9.1–14.5) | 14.8 (11.5–21.7) | 0.097 |
| Lymphocytes, cells/μL | n.a. | 1,288 (939–1.579) | 1,035 (284–1.706) | 0.521 |
| Monocytes, cells/μL | n.a. | 495 (448–742) | 738 (599–1.005) | **0.009** |
| Platelet count, x 1000/μL | n.a. | 198 (154–324) | 187 (131–240) | 0.125 |

Continuous variables are represented as median and interquartile range. Categorical variables are represented as *n* (frequency %).
n.a. – not applicable
<sup>1</sup>Coronary artery disease or congestive heart failure.
Categorical variables were compared using the two tailed Fisher exact test, and the continuous variables using Student's *t* or ANOVA tests for parametric and the Mann Whitney U or Kruskal-Wallis tests for nonparametric distributions. Significant *p* values are in bold.

#### Sample preparation

A total of 30 μL of plasma (in duplicate) were mixed with the same volume of the IS mixture and 500 μL of a degassed mixture of pre-chilled acetonitrile/isopropanol/water (3:3:2, v/v/v). After vortexing for 20 s and incubating in ice in ultrasonic water bath for 5 min, samples were centrifuged at 12,000 × *g* at 4°C for 5 min and the supernatant (480 μL) was dried under Nitrogen gas. Samples were reconstituted with 60 μL of acetonitrile/water (1:1, v/v) containing 2 μg mL^−1^ of the IS *p*-fluoro-L-phenylalanine, vortexed for 15 s, and centrifuged as above. The resulting supernatant was used for LC-MS analysis.

A pooled quality control (QC) sample was prepared combining 5 μL of each plasma before extraction and processed in the same way as the specimen samples. QC samples were injected every 10^th^ biological sample to monitor the stability of the analytical system as well as the reproducibility of the procedure for sample treatment [36].

Subgroups of pooled QC samples (control, survivors, or non-survivors) were created to collect fragmentation spectra via data-dependent acquisition (DDA) mode on the mass spectrometer. The analysts running MS-based experiments were blinded on the sample grouping until the end of data analysis to limit biased peak annotation.

#### LC-MS conditions

Liquid chromatography (LC) analysis was performed on a Dionex UltiMate 3000 UHPLC (Thermo Fisher Scientific, Bremen, Germany) system using a Waters^®^ ACQUITY UPLC^®^ BEH amide column (150 x 2.1 mm x 1.7 μm) by gradient elution at a constant flow rate of 350 μL min^−1^. The column oven temperature and injection volume were set to 40°C and 5.0 μL, respectively.

Two different mobile phase compositions, with different pH values, were used. One consisted of (A) water:acetonitrile (95:5, v/v) and (B) acetonitrile:water (95:5, v/v) both with 0.1 % formic acid (pH 3) and the other consisted of (A) water:acetonitrile (95:5, v/v) and (B) acetonitrile:water (95:5, v/v) both with 0.05 % ammonium hydroxide and 10 mM ammonium acetate (pH 8). The gradient elution was 0-0.5 min 100 % B; 0.5-5.0 min 45 % B; 5.0-9.0 min 45 % B; 9.0-10.0 min 100 % B; 10.0-15.0 min 100 % B.

The LC was coupled to a hybrid Quadrupole-Orbitrap high resolution and accurate mass spectrometer (QExactive Plus, Thermo Scientific) equipped with a heated electrospray ion source operating in both negative (ESI-) and positive (ESI+) ionization modes. Source ionization parameters were: spray voltage −3.6 kV/+3.9 kV; capillary temperature 270°C; probe heater temperature 380°C; S-Lens RF level 50, sheath and auxiliary gases 50 and 10 (arbitrary units), respectively. Samples were analyzed in Full MS mode in the scan range of *m/z* 50-710 at the resolution of 70,000 FWHM (full width at half maximum). The automatic gain control (AGC) target was set at 1.0e6 with a maximum injection time (IT) of 150 ms.

The solution of target analytes and the subgroups pooled QC samples were analyzed in Full MS followed by data-dependent acquisition (dd-MS^2^ top5 experiment) in the same scan range as above. For the full MS scan, the mass resolution was set to 17,500 FWHM with the following settings: AGC target 1.0e6 and maximum IT of 80 ms. For the dd-MS^2^ scan, the mass resolution was set to 17,500 FWHM with following settings: AGC target at 1.0e5, maximum IT of 50 ms, isolation window at *m/z* 1.2, normalized collision energy (NCE) 15, 35 (ESI+) and 10, 30 (ESI-), intensity threshold at 1.0e6, exclude isotopes "on", and dynamic exclusion of 10.0 s.

#### Non-targeted and targeted LC-HRMS-based metabolomics

For the non-targeted analysis, the LC-HRMS data files were submitted to a metabolomics workflow using MS-DIAL software (RIKEN, version 4.80) [37] for data processing, including peak matching against an MS/MS library. The parameters used for both pH 8 and pH 3 analyses are described in Supplementary table 3. Features were selected assuming coefficient variation (CV) % values less than 30 % in QC samples and Gaussian-like peak shape according to the protocols for quality control used in untargeted metabolomics [36, 38].

Compound annotation was done i) based on the MS/MS fragment comparison with the standard compounds, ii) by comparing the aligned *m/z* ions with a mass error below 6 ppm to those available at the HMDB [34] and METLIN website, and iii) by comparing the investigated MS/MS spectra with a similarity score ≥ 80% to those in the NIST 20 Tandem Mass Spectral Library and MassBank of North America using a customized MSP file in MS-DIAL. Furthermore, molecular formulas were determined using MS-FINDER (RIKEN, version 3.52) [39].

Targeted data analysis was performed as a confirmation step for the non-targeted approach, in TraceFinder software v3.1 (ThermoFisher Scientific, Waltham, Massachusetts, EUA). An in-house library that includes retention time, exact mass and fragments of the target compounds was used. As identification criteria, mass errors less than 5 ppm and retention time variation of <1 % compared to the defined retention time were accepted [40]. Supplementary Tables 1 and 2 present the target compounds monitored via the method at pH 8 and pH 3, respectively.

### Statistical Analysis

Sample size was determined by the feasibility of recruitment and eligibility criteria.

Data distribution was analyzed using the Shapiro-Wilk test and median and interquartile intervals, and the Mann–Whitney or Kruskal-Wallis tests were used for data with asymmetrical distribution. Categorical variables were compared using the Fisher’s exact test with absolute (n) and relative (%) frequencies.

Data derived from processed NMR spectra was subjected to the multivariate statistics Principal Component Analysis using the MetaboAnalyst 4.0 [41] platform. For the univariate statistics, Kruskal-Wallis and Dunn’s post-hoc tests were used for variables’ comparison of non-transformed NMR or MS data. Interaction between sex and disease was analyzed using two-way ANOVA and Tukey’s multiple comparison tests. GraphPad Prism version 8.4.3 was used for all analysis. *P* < 0.05 was considered for rejection of the null hypothesis.

Classification and regression tree (CART) models [42] were fitted to assess which metabolites best predict the observed morbidity class of study participants. The model fitting algorithms have been implemented in library "rpart" [43] for the R programming language. We set “method = “class” and the remaining parameters were kept at the default values for all models. For CART models, the results of NMR-based metabolomics were used, in addition to the subjects’ sex and age.

## Results

### Subjects’ demographics and clinical parameters

A total of 47 plasma samples were included in this study; 12 samples were from non-infected control subjects and 35 samples from individuals with severe cases of COVID-19, grouped into survivors (n= 18) and non-survivors (n= 17), according to the 28-day mortality outcome. The demographics and clinical characteristics of all subjects included in the study are shown in Table 1. Briefly, age, sex distribution and co-morbidities were similar among groups. The non-invasive oxygen supplementation was used in 44 % of subjects in the survivors group whereas 100 % of the subjects in the non-survivors group received mechanical ventilation.

Laboratory findings revealed that patients in the survivors group presented ∼ 50 % more monocyte counts (*p*=0.009) if compared to non-survivors. At admission in the ICU, leukocytes and platelet counts were similar between the two groups of infected patients.

### ^1^H NMR- and MS-based metabolomics

Representative spectra of aliphatic (Figs 1A and 1B), amidic and aromatic (Fig 1C) regions indicate important differences in the metabolite profile among the three groups. Discriminating metabolites as (CH_3_)_3_ choline-related metabolites, creatine/creatinine, amino acids, organic acids, and broad residual signals of lipids are depicted followed by arrows indicating higher/lower contents in severe COVID-19 cases. A discriminating metabolite profile was confirmed with Principal Component Analysis (PCA) scores plot (Fig 1D) where principal components 1, 2, and 3 accounted for approximately 70 % of the variation among groups. PCA loading factors plot highlights (CH3)_3_ choline, creatine/creatinine, lactate, acetate, and broad signals of CH_3_ and CH_2_ lipoproteins as discriminating variables (Supplementary Fig 1).

**Figure 1.**
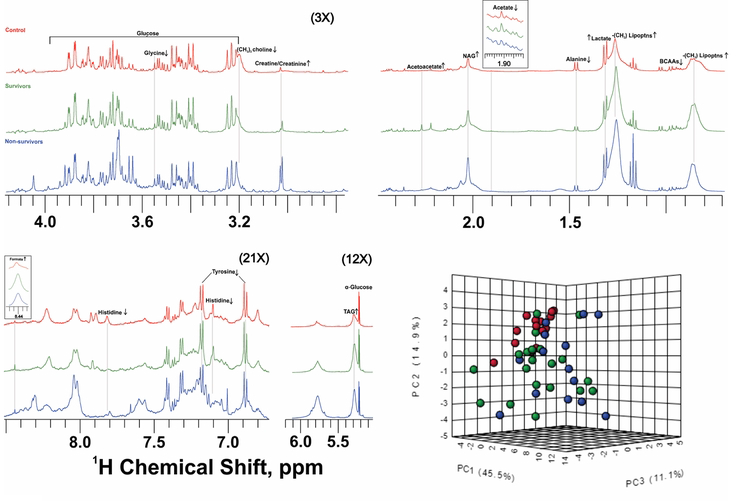
^1^H NMR-based metabolomics shows different plasma metabolite profiling in severe COVID-19 patients compared to control subjects. ^1^H NMR representative spectra of control (red), COVID-19 survivors (green) and non-survivors (blue). Metabolites that differ significantly among groups are indicated as higher (↑) or lower (↓) contents compared to controls; (A) and (B) aliphatic region (3 × magnified); (C) amidic and aromatic regions (21 × magnified); (D) Principal Component Analysis 3D scores plot shows discriminating profiling among groups.

To gain meaningful insight into the changes associated with disease severity and outcome, metabolites that exhibited significant differences in content among groups, according to the PCA results, were selected. NMR-based metabolomics revealed that the levels of (CH_3_)_3_-choline metabolites, including glycerophosphocholine, phosphocholine, and choline, as well as glycine, which is associated with one-carbon metabolism, were significantly lower in both survivors and non-survivors, if compared to controls (Figs 2A and 2B). Additionally, ^1^H NMR-based metabolomics identified several metabolites related to glucose, insulin sensitivity, and inflammation that were significantly higher in infected subjects (Figs 2C-G), including creatine/creatinine (considered together due to signal overlap), *N*-acetyl ^1^H of glycoproteins, and lactate. Importantly, creatine/creatinine set apart non-survivors from survivors and control subjects, as the levels were higher in the former. Lower levels of acetate and higher levels of formate, a byproduct of bacteria metabolism in the gut, were observed in infected subjects but not in controls (Figs 2F and G). Additionally, residual signals of (CH_2_) of VLDL-lipoproteins, lipids (CH=CH olefinic protons of triacylglycerols) and acetoacetate, a ketone body, were significantly higher in both groups of infected subjects if compared to controls (Figs 2H-J).

**Figure 2.**
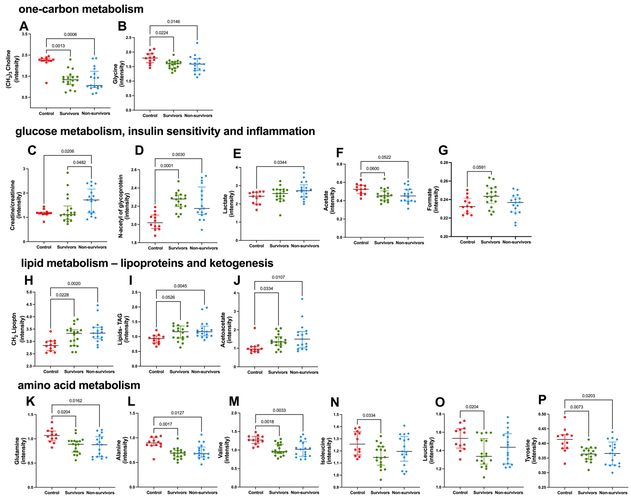
Metabolites that were significantly altered in severe COVID-19, according to ^1^H NMR-based metabolomics. Most discriminating metabolites according to PCA loading factors, presenting significant differences among groups. Control, n= 12 (red circles); Survivors, n=18 (green circles); Non-survivors, n=17 (blue circles). Metabolites related to one-carbon metabolism (A,B): (CH_3_)_3_ choline-related metabolites and glycine; metabolites related to glucose metabolism, insulin sensitivity and inflammation (C-G): creatine/creatinine, *N*-Acetyl of glycoproteins, lactate, acetate and formate; metabolites related to lipid metabolism (H-J): CH_2_ lipoproteins (mainly VLDL), lipids-TAG (CH=CH olefinic protons of triacylglycerols) and acetoacetate; metabolites related to amino acids and protein metabolism (K-P): glutamine, alanine, valine, isoleucine, leucine, tyrosine; metabolites’ contents were determined according to their respective peak intensity. Data were presented as median with interquartile range and only significant *P* values are shown, according to Kruskal-Wallis and Dunn’s post-hoc tests.

^1^H NMR-metabolomics findings also suggest that the dysregulation in amino acid metabolism is a function of severe COVID-19, as survivors and non-survivors had plasma levels of glutamine, alanine, branched-chain amino acids (valine, leucine, isoleucine) and tyrosine lower than those observed in controls (Figs 2K-P). Supplementary Tables 4 and 5 present the ^1^H NMR assignment information for the metabolites and broad signals of lipids and proteins that distinguished the groups, in both PCA and univariate analysis.

To further investigate the changes in plasma metabolome associated with severe COVID-19, LC-high-resolution mass spectrometry (LC-HRMS)-based metabolomics was used. In the current study, LC-HRMS allowed us to confirm the alterations detected by the ^1^H NMR-based metabolomics in amino acids and protein metabolism and in ketogenesis. Importantly, it allowed us to discriminate the metabolic abnormalities associated with fatal COVID-19 and to complement the NMR data, providing insightful information regarding the alterations in one-carbon metabolism and in metabolites associated with insulin sensitivity and inflammation. Metabolites related to one-carbon metabolism glycerophosphocholine, serine, betaine, and histidine were lower (Figs 3A-D) whereas xanthine and hypoxanthine were higher in infected subjects, if compared to controls (Figs 3E and 3F). The lower levels of glycerophosphocholine in the survivors and non-survivors groups, when compared to control subjects, support the ^1^H NMR-based metabolomics results, which detected lower (CH_3_)_3_ choline-related metabolites in the two groups of infected patients. Metabolites associated with inflammation were significantly higher in infected subjects, if compared to controls. Importantly, creatinine, 4-hydroxyproline, gluconic acid and *N*-acetylserine were significantly higher in the non-survivors group, when compared to both survivors and control groups (Figs 3G-J). The higher levels of creatinine in non-survivors confirmed the results of the NMR analysis (Fig 2C). Moreover, asymmetric dimethylarginine and methylmalonic acid were significantly higher in survivors and non-survivors, when compared to control subjects (Figs 3K and 3L). MS-based metabolomics revealed significantly higher content of β-hydroxybutyrate – a ketone body (Fig 3M) and lower content of the amino acid tryptophan (Fig 3N) in infected subjects, if compared to controls. Supplementary Fig 2 shows the essential stability of the QC samples throughout the run, which was within SD limit. Additionally, the IS *p*-fluoro-DL-phenylalanine and U-^13^C D-glucose, which were added to all samples, showed that the CV was less than 10 %, indicating that the differences observed in metabolites among groups could be attributed to biological variation and not to the variability of the analytical system. Supplementary Tables 6 and 7 show the parameters of the assigned metabolites with significant differences among groups according to the non-targeted MS-metabolomics in the negative and positive modes, respectively.

**Figure 3.**
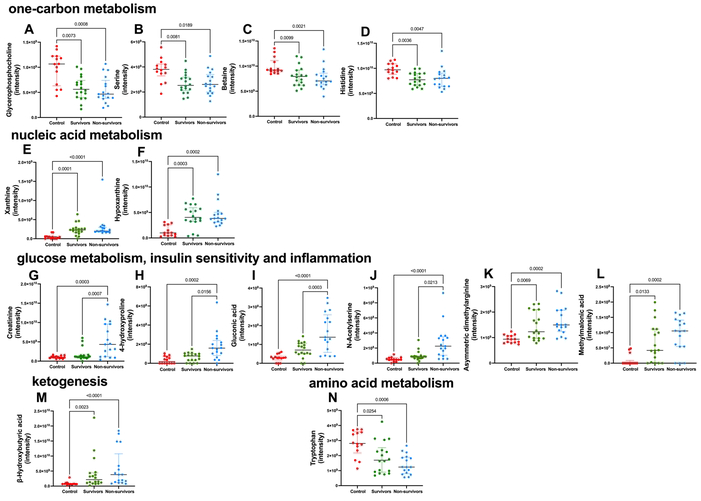
High-resolution mass spectrometry-based metabolomics shows altered plasma metabolic profile in severe COVID-19. Assigned metabolites in the untargeted and confirmed in the targeted approach, presenting significant differences among groups. Control, n=12 (red circles); Survivors, n=18 (green circles); Non-survivors, n=17 (blue circles). Metabolites related to one-carbon metabolism (A-D): glycerophosphocholine, serine, betaine, histidine and to nucleic acid metabolism (E-F): xanthine, hypoxanthine; metabolites related to glucose metabolism, insulin sensitivity and inflammation (G-L): creatinine, 4-hydroxyproline, asymmetric dimethylarginine, gluconic acid, N-acetyl serine, methylmalonic acid. Metabolite related to lipid metabolism (M): β-hydroxybutyrate; metabolite related to amino acid metabolism (N): tryptophan. Metabolites’ contents were determined according to their respective peak intensity. Data were presented as median with interquartile range and only significant *P* values are shown, according to Kruskal-Wallis and Dunn’s post-hoc tests.

CART models were built to assess the predictive power of metabolites levels in classifying study participants into control, survivors and non-survivors. This model was built using the data from the NMR-metabolomics and considered only the metabolites that showed discriminatory power among the groups identified by the multivariate PCA analysis (Fig 2). Additionally, the subjects’ sex and age were included in the model. Supplementary Figure 3 shows a CART model fitted to data indicating that choline-related metabolites had the highest predictive power with a first partition point at 2.064. The classification tree shows that over 92 % of control subjects followed the classification of ≥2.064, and most survivors (95 %) and non-survivors (89%) were classified as < 2.064. In addition to the first partition point at choline-related metabolites, we also observed a second partition criteria at creatine/creatinine < 1.473. For this second partition, most survivors were included (∼70 %). Conversely, for the non-survivors, the majority were classified at ≥ 1.473 (64 %). Supplementary Fig 3 presents the final allocation contingent among the morbidity classes achieved by applying the two classification criteria. These results clearly suggest that choline-related metabolites have a protective effect. As opposed to creatine/creatinine, where higher contents predicted fatal outcomes.

### Lipoproteins dynamics

A complete characterization of lipoproteins was performed (Fig 4) and significant changes were observed as a function of severe COVID-19. The results show that survivors and non-survivors presented lower concentrations of total cholesterol, LDL, and HDL, if compared to controls (Figs 4A-C). VLDL and triacylglycerol concentrations were significantly higher (Figs 4D-E) and non-HDL cholesterol (Fig 4E) was significantly lower in the non-survivors, if compared to controls.

**Figure 4.**
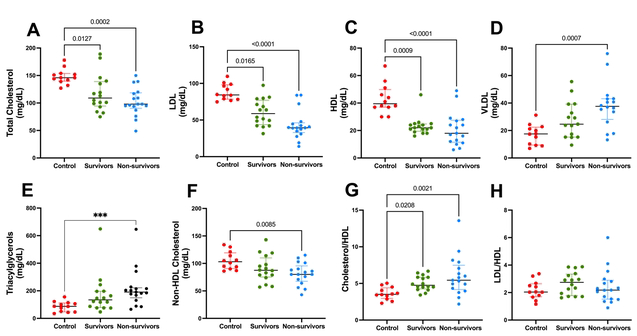
Lipoprotein dynamics changed significantly in severe COVID-19. (A) Total Cholesterol, (B) Low Density Lipoprotein - LDL, (C) High Density Lipoprotein - HDL, (D) Very Low-Density Protein - VLDL, (E) Triacylglycerol, (F) non-HDL cholesterol, (G) Cholesterol-to-HDL ratio and (H) LDL-to-HDL ratio. Control, n= 12 (red circle); Survivors, n=18 (green circle); Non-survivors, n=17 (blue circle). Data were presented as median with interquartile range and only significant *P* values are shown, according to Kruskal-Wallis and Dunn’s post-hoc tests.

### Sex-based differences in lipoproteins and metabolites

Lastly, we investigated whether changes in lipoproteins and in metabolites associated with severe COVID-19 differed according to sex, by two-factor analysis (Fig 5). In general, the changes observed in female subjects mirrored the changes described above, when considering all subjects. Our findings indicate that the strongest effect was seen in severe COVID-19 cases and no differences between men and women within group were observed. However, for HDL, total cholesterol, non-HDL cholesterol, LDL to HDL and HDL to cholesterol ratios, significant interactions between disease and sex were observed. Additionally, lower contents of HDL (Fig 5C) and higher contents of VLDL (Fig 5D) and triacylglycerols (Fig 5E) were observed in female but not in male subjects in the non-survivors group, when compared to controls.

**Figure 5.**
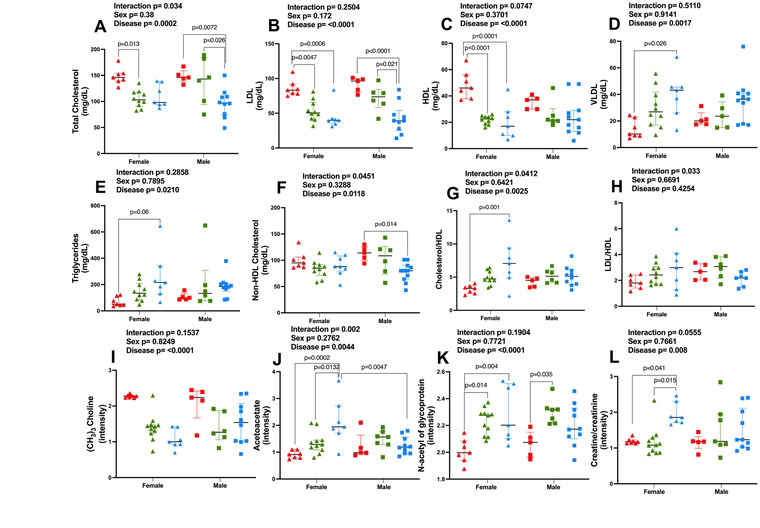
Severe COVID-19 induced changes in lipoproteins and metabolites that are greater in women than in men. (A) Total Cholesterol, (B) Low Density Lipoprotein - LDL, (C) High Density Lipoprotein - HDL, (D) Very Low-Density Protein - VLDL, (E) Triacylglycerol, (F) non-HDL cholesterol, (G) Cholesterol-to-HDL ratio and (H) LDL-to-HDL ratio, (I) (CH_3_)_3_ Choline, (J) Acetoacetate, (K) N-acetyl of glycoprotein, (L) Creatine/creatinine. Female subjects: control, n=7 (red triangles); survivors, n=11 (green triangles); non-survivors, n=7 (blue triangles). Male subjects: control, n= 5 (red squares); survivors, n=7 (green squares); non-survivors, n=10 (blue squares). Data were presented as median with interquartile range and *P* values are shown, according to two-factor ANOVA and Tukey’s multiple comparison tests.

The same sex-related pattern changes were observed for selected metabolites analyzed by ^1^H NMR-based metabolomics, where the strongest effect was seen in severe COVID-19 cases (Figs 5I-L). Lower content of (CH_3_)_3_ choline-related metabolites (Fig 5I) and higher content of *N*-acetyl of glycoproteins (Fig 5K) were observed among females in both survivors and non-survivors, if compared to controls, whereas among men these differences were observed only between survivors and controls. Additionally, higher contents of acetoacetate (Fig 5J) and creatine/creatinine (Fig 5L), as a function of severe COVID-19, were only observed in women but not in men. Indeed, for acetoacetate and creatine/creatinine, significant interactions between disease and sex were observed. There were no significant differences in amino acids and one-carbon metabolism-related compounds when sex was considered as a variable.

The metabolic alterations observed in severe COVID-19 are shown in Fig 6.

**Figure 6.**
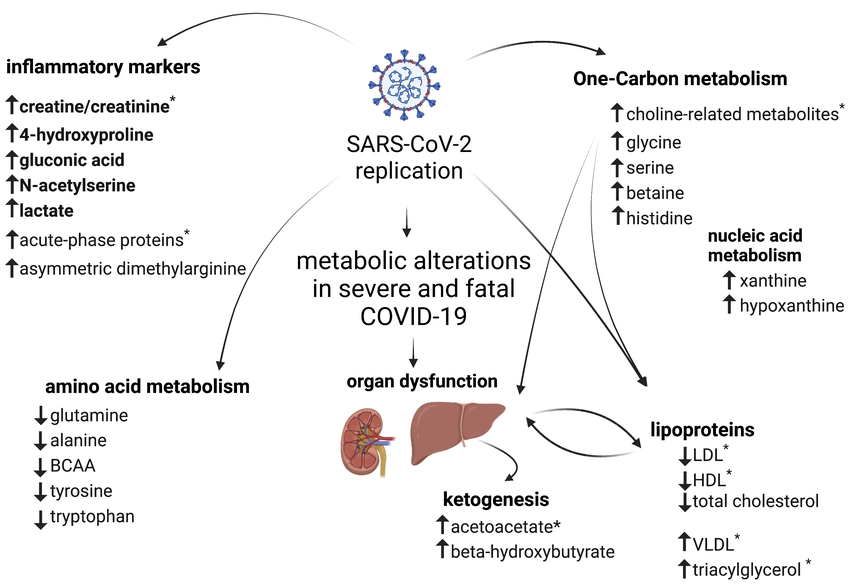
Metabolic alterations in severe COVID-19. Summary of changes in plasma metabolites in survivors and non-survivors subjects. Metabolites that differ significantly among groups are indicated as higher (↑) or lower (↓) contents compared to controls. Metabolites in bold were higher in the fatal cases of the disease compared to survivors and controls. * indicates metabolites with changes greater in women than in men. This figure was created with BioRender.com.

## Discussion

The present study aimed to investigate plasma metabolic changes in samples from survivors and non-survivors of severe COVID-19 infection. Our goal was to search for potential metabolic pathways that could be involved in COVID-19 pathophysiology and disease outcome. We identified significant changes in a plethora of metabolites, indicating that severe COVID-19 dysregulates one-carbon, lipid, and amino acid metabolism and lipoprotein dynamics.

Higher contents of creatine/creatinine, 4-hydroxyproline, gluconic acid and *N*-acetylserine in non-survivors, if compared with survivors and control groups, indicate that these metabolites are associated with uncontrolled inflammation, multi-organ dysfunction, particularly liver and kidneys, and some degree of insulin resistance in fatal COVID-19. They may be considered as biomarkers for prognostic purposes and to monitor how different organs and tissues respond to the infection. For instance, gluconic acid has been previously linked to hyperglycemia and brain injury in ischemic stroke [44], and it may be considered a marker of oxidative stress. Additionally, *N*-acetylation of amino acids, including the formation of *N*-acetylserine, has been associated with SARS-CoV-2 infection and COVID-19 pathogenesis [45] whereas higher levels of *N*-acetylserine have been recently considered a marker of progression of chronic kidney disease [46].

Higher plasma contents of creatine/creatinine can be an indication of lower sensitivity to insulin, as we observed in the fatal cases of COVID-19. Although higher creatine levels in severe COVID-19 have been associated with kidney dysfunction [26], and are important for viral replication [47], recent findings place creatine as a key metabolite involved in the regulation of adipocyte thermogenesis and whole-body energy metabolism and immunity [48,49]. Therefore, higher levels of creatine/creatinine in infected subjects can be regarded as a biomarker of detrimental effects on metabolic health and immune responses imposed by SARS-CoV-2 infection.

Higher levels of 4-hydroxyproline found in non-survivors is a strong indication of disturbed amino acid metabolism induced by SARS-CoV-2 infection, as already suggested [20, 23-25]. In humans, 4-hydroxyproline in blood is a product of protein degradation, mainly collagen. Most 4-hydroxyproline is recycled back to synthesize glycine by the liver and kidneys and this seems to be an important source of glycine for cells and tissues [50]. Therefore, the significantly higher levels of 4-hydroxyproline in non-survivors can be regarded as a feature of severe COVID-19, suggestive of liver dysfunction and lower availability of glycine. This result reveals a potential disruption in the one-carbon metabolism pathway, which is significant as glycine serves as a crucial methyl donor in this pathway. This disruption has already been described previously [27]. In addition to glycine, significant lower contents of serine, betaine and histidine, which are metabolites also involved in the one-carbon metabolism, were found in infected subjects. Importantly, the significant drop in choline-related metabolites (as observed in the NMR-based metabolomics) which agrees with a drop in glycerophosphocholine (by MS-based metabolomics), strengths the idea that severe COVID-19 alters one-carbon metabolism and affects the availability of methyl donors. Indeed, a comprehensive characterization of the alterations in one-carbon metabolism would help integrating the metabolic changes in glucose, amino acid, nucleotide, and lipid metabolism [51], as well as provide us with a better understanding of the alterations in epigenetic regulation and redox homeostasis associated with severe COVID-19.

Phosphatidylcholine is the most abundant phospholipid and in humans its synthesis is probably the main point of deviation for one-carbon donors [52]. Our results show that severe COVID-19 alters lipoprotein dynamics, as indicated by lower contents of total cholesterol and HDL- and LDL-cholesterol and higher contents of VLDL and triacylglycerols, particularly in fatal COVID-19 (Fig 4). Therefore, one may speculate that the alterations in one-carbon metabolism observed in infected subjects caused the drop in choline-related metabolites which significantly disrupted lipoproteins dynamics, as indicated by other studies [52,53]. Conversely, lower choline availability may have contributed to reduced phospholipid synthesis by the liver and disturbed lipoproteins dynamics, as choline deficiency is known to alter lipid metabolism and to induce liver fibrosis [53]. Another indication of the effect of SARS-CoV-2 infection on host lipid metabolism and lipoproteins dynamics was recently shown by our group, where simvastatin, by disrupting lipid rafts in human epithelial lung cells, prevented SARS-CoV-2 entry and replication [54]. Importantly, NMR-based metabolomics has also shown that alterations in lipoprotein dynamics may be associated with systemic effects of COVID-19, even in subjects in the recovery phase of the disease. Particularly, the HDL subfraction negatively correlates with inflammatory cytokines [55]. Additionally, apolipoproteins alterations are more pronounced in the fatal cases of COVID-19 [56].

This scenario is compatible with the CART model fitted to data (Supplementary Fig 3), which indicates that choline-related metabolites had the highest predictive power with a first partition point at 2.064, demonstrating that most survivors (95 %) and non-survivors (89%) were classified as < 2.064; and that higher content of creatine/creatinine is a strong predictor of disease fatality. Recent evidence links choline metabolism in the liver and the gut microbiota to endothelial function and thrombosis [57,58], which is one of the clinical manifestations associated with disease progression and severe COVID-19 [59]. Indeed, choline seems to be essential to sustain mitochondrial energetics and maximal platelet activation, and, therefore, to regulate thrombosis [60].

We also found increased levels of asymmetric dimethylarginine in infected subjects, which is produced by arginine methylation. Therefore, asymmetric dimethylarginine synthesis is directly related to one-carbon metabolism for methyl donor availability. Additionally, asymmetric dimethylarginine has been strongly associated with fibrosis in the liver, kidney, and heart [61,62], tissues that are compromised in COVID-19. Therefore, these data strongly support that alterations in one-carbon and amino acid metabolism are linked to liver damage in most severe outcomes of COVID-19.

Our study confirmed that severe COVID-19 induced important changes in amino acid metabolism, where significantly lower contents of alanine, BCAA (valine, isoleucine, leucine), glutamine, tryptophan, glycine, and tyrosine were found in both survivors and non-survivors, if compared to control subjects. Lower amino acid levels may indicate an increase in amino acid catabolism to provide the necessary supply of carbons and ATP to support viral replication [16, 63, 64]. For instance, isoleucine [65] and glutamine [66] seem to be essential for SARS-CoV-2 replication. In the case of glutamine, it has been shown that inhibition of glutaminolysis halted SARS-CoV-2 in primary astrocytes in a rodent model; authors suggested that lower availability of glutamine is a contributing factor for the neurological impairments observed in long post-COVID. Additionally, a decrease in plasma glutamine [67] and lower salivary levels of tyrosine and BCAA [68] seem to be associated with COVID-19 severity, which supports our findings.

Lower levels of glutamine in the plasma may indicate an increased use of this amino acid, especially by the host’s liver and immune cells. This increased use may be needed to support the synthesis of proteins and inflammatory mediators during the acute phase of infection. Indeed, the liver is actively involved in the synthesis of acute-phase proteins. In this context, the assigned broad signals of *N*-Acetyl of glycoproteins, which were higher in severe COVID-19 subjects (Fig 2D), may include signals from the acute-phase proteins, such as α_1_-acid glycoprotein, α_1_-antitrypsin, and haptoglobin and to the ^1^H from sidechains of N-acetyl-glucosamine and N-acetylneuraminic acid [69]. ^1^H signals of *N*-acetyl of glycoproteins have been suggested to be markers of SARS-CoV-2 infection and inflammation and to be implicated in long post-COVID symptoms as well [70]. Indeed, enrichment analysis of genes associated with cases of severe COVID-19 indicates acute phase response and inflammation among the most significant biological functions altered, accordingly to a recently multi-omic analysis of COVID-19 datasets [71].

Other authors have shown that liver infection by SARS-CoV-2 directly contributes to hepatic dysfunction and that deceased subjects often presented abnormal liver enzymes, microvesicular steatosis and mild inflammatory infiltrates in the hepatic lobule and portal tract [72,73]. The increase in purine metabolites (xanthine and hypoxanthine) may also be regarded as a direct effect of SARS-CoV-2 infection in the liver that enables virus replication. Higher levels of deoxycytidine, associated with increased viral load, have already been associated with severe and fatal outcomes [74]. Accordingly, the liver of patients with severe COVID-19 shares many similarities with the non-alcoholic fatty liver disease (NAFLD), a common manifestation of the metabolic syndrome that can progress to hepatocyte injury, inflammation, and fibrosis [75,76].

Lastly, the higher levels of acetoacetate and ß-hydroxybutyric acid observed in infected subjects also reflect the impact of severe COVID-19 on liver function. Dysregulation in ketogenesis is also associated with the pathogenesis of NAFLD and decreased insulin sensitivity, an important manifestation of metabolic syndrome. In this scenario, enhanced ketogenesis seems to be a consequence of increased influx and accumulation of lipids in the liver (e.g., triacylglycerols), which in turn results in an increased flux of Acetyl-CoA [77].

This is the first study to demonstrate host metabolic disturbances associated with severe COVID-19 and to investigate the responses in the fatal cases of the disease in hospitalized subjects in Rio de Janeiro, during the early months of the COVID-19 pandemic in Brazil. Whilst our study provides new insights into the metabolic disturbances associated with fatal outcomes of COVID-19, one limitation is its reduced sample size. Nonetheless, since the incidence of severe outcomes after hospital discharge can be very high in Brazil [73], these metabolic alterations may be considered to improve our understanding of the pathophysiology of long-post COVID.

Our results show that alterations in key metabolites, such as choline-metabolites, creatinine/creatinine, ^1^H signals of N-acetyl of glycoproteins, and acetoacetate, as well as changes in lipoproteins are greater in women. Even though current evidence indicates that men are more vulnerable to severe COVID-19 and higher mortality rates have been observed in this category [78], women seem to be more susceptible to developing long COVID syndrome [79]. Our study highlights the importance of considering the sex-based differences here described, when monitoring and treating COVID-19 patients. These differences should also be considered when designing strategies for pandemic surveillance. Doing so may lead to better disease stratification and improved patient outcomes.

## Data Availability

All relevant data are within the manuscript and its Supporting Information files.

## Acknowledgments

The authors acknowledge the contribution of the subjects who participated in this study, without receiving any direct compensation.

## Supporting Information Caption

**Supplementary Table 1**: List of the target compounds monitored in the method at pH8.

**Supplementary Table 2**: List of the target compounds monitored in the method at pH3.

**Supplementary Table 3**: Parameters used for untargeted analysis in MS-DIAL software.

**Supplementary Table 4**: Assignment table of metabolites that discriminated in the PCA and univariate analyses.

**Supplementary Table 5**: Assigned metabolites with significant differences among groups according to the non-targeted MS-metabolomics in the negative mode.

**Supplementary Table 6**: Assigned metabolites according to the non-targeted MS-metabolomics in the positive mode.

**Supplementary Figure 1**. PCA loading factors plot highlights discriminating metabolites according to ^1^H NMR-based metabolomics.

**Supplementary Figure 2**. PC1 scores plot versus sample in run order at pH 8 and pH 3 analyses indicating the stability of the system throughout the analytical run.

**Supplementary Figure 3: Classification and Regression Tree (CART) model indicates that NH3-choline related metabolites and creatine/creatinine present high predictive power in assigning subjects according to their morbidity class.** The CART model was built using the metabolites that were found to be discriminatory in the multivariate PCA analysis, which was based on ^1^H NMR metabolomics: NH_3_-choline-related metabolites, creatine/creatinine, glutamine, alanine, histidine, tyrosine, valine, leucine, isoleucine, CH2 of lipoproteins (VLDL), CH=CH olefinic protons of triacylglycerols, acetoacetate, lactate, acetate, formate, *N*-acetyl of glycoproteins. In red is the classification that best predicted the morbidity class for control subjects (partition point choline ≥2.064), survivors-creatine/creatinine < 1.473 and non-survivors-creatine/creatinine ≥ 1.473.

## References

1. Cohen LE, Spiro DJ, Viboud C. Projecting the SARS-CoV-2 transition from pandemicity to endemicity: Epidemiological and immunological considerations. PLoS Pathog. 2022; 30;18(6):e1010591. doi: 10.1371/journal.ppat.1010591.

2. WHO. COVID-19 Weekly Epidemiological Update 35. World Health Organization. 2021 Dec:1–3.

3. Brasil. Secretarias Estaduais de Saúde. Painel Coronavirus. 2023. Available from: https://covid.saude.gov.br/2023

4. Raoult D, Zumla A, Locatelli F, Ippolito G, Kroemer G. Coronavirus infections: Epidemiological, clinical and immunological features and hypotheses. Cell Stress. 2020, 4: 66–75. doi: 10.15698/cst2020.04.216

5. Xiao N, Nie M, Pang H, Wang B, Hu J, Meng, X. et al. Integrated cytokine and metabolite analysis reveals immunometabolic reprogramming in COVID-19 patients with therapeutic implications. Nat Commun. 2021; 12: 1618. doi: 10.1038/s41467-021-21907-9

6. WHO. Coronavirus disease (COVID-19): Post COVID-19 condition. World Health Organization. 2021. Available from: https://www.who.int/news-room/questions-and-answers/item/coronavirus-disease-(covid-19)-post-covid-19-condition

7. Carfì A, Bernabei R, Landi F, Gemelli Against COVID-19 Post-Acute Care Study Group. Persistent Symptoms in Patients After Acute COVID-19. JAMA. 2020, 324: 603–605. doi: 10.1001/jama.2020.12603.

8. Fernández-de-Las-Peñas C, Rodríguez-Jiménez J, Cancela-Cilleruelo I, Guerrero-Peral A, Martín-Guerrero JD, García-Azorín D, et al. Post-COVID-19 Symptoms 2 Years After SARS-CoV-2 Infection Among Hospitalized vs Nonhospitalized Patients. JAMA Netw Open. 2022; 5(11):e2242106. doi: 10.1001/jamanetworkopen.2022.42106.]

9. dos Santos GC, Renovato-Martins M, de Brito NM. The remodel of the “central dogma”: a metabolomics interaction perspective. Metabolomics. 2021; 17 (5):48. doi: 10.1007/s11306-021-01800-8.

10. Byers NM, Fleshman AC, Perera R, Molins CR. Metabolomic insights into human arboviral infections: Dengue, chikungunya, and zika viruses. Viruses. 2019; 11 (3): 225. doi:10.3390/v11030225

11. El-Bacha T, Struchiner CJ, Cordeiro MT, Almeida FCL, Marques ET, Da Poian AT. ^1^H Nuclear Magnetic Resonance Metabolomics of Plasma Unveils Liver Dysfunction in Dengue Patients. J Virol. 2016; 90(16): 7429–7443. doi: 10.1128/JVI.00187-16

12. Wu Q, Zhou L, Sun X, Yan Z, Hu C, Wu J, et al. Altered Lipid Metabolism in Recovered SARS Patients Twelve Years after Infection. Sci Rep. 2017; 7(1):9110. doi: 10.1038/s41598-017-09536-z.

13. Melo CFOR, Delafiori J, de Oliveira DN, Guerreiro TM, Esteves CZ, Lima E de O., et al. Serum metabolic alterations upon ZIKA infection. Front Microbiol. 2017; 8: 1954. doi: 10.3389/fmicb.2017.01954

14. Diop F, Vial T, Ferraris P, Wichit S, Bengue M, Hamel R, et al. Zika virus infection modulates the metabolomic profile of microglial cells. PLoS One. 2018; 13: e0206093. doi.org/10.1371/journal.pone.0206093

15. Girdhar K, Powis A, Raisingani A, Chrudinová M, Huang R, Tran T, et al. Viruses and Metabolism: The Effects of Viral Infections and Viral Insulins on Host Metabolism. Annu Rev Virol. 2021; 8(1): 373–391. doi: 10.1146/annurev-virology-091919-102416.

16. El-Bacha, T.; Da Poian, A.T. Virus-induced changes in mitochondrial bioenergetics as potential targets for therapy. Int J Biochem Cell Biol. 2013; 45 (1): 41–46. doi: 10.1016/j.biocel.2012.09.021. Epub 2012 Oct 2.

17. Wolff G, Limpens RWAL, Zevenhoven-Dobbe JC, Laugks U, Zheng S, de Jong AWM. et al. A molecular pore spans the double membrane of the coronavirus replication organelle. Science. 2020; 369 (6509): 1395–1398. doi: 10.1126/science.abd3629.

18. Dias SSG, Soares VC, Ferreira AC, Sacramento CQ, Fintelman-Rodrigues N, Temerozo JR, et al. Lipid droplets fuel SARS-CoV-2 replication and production of inflammatory mediators. PLoS Pathog. 2020; 16 (12): e1009127. doi: 10.1371/journal.ppat.1009127

19. Ricciardi S, Guarino AM, Giaquinto L, Polishchuk EV, Santoro M, Di Tullio G, et al. The role of NSP6 in the biogenesis of the SARS-CoV-2 replication organelle. Nature. 2022; 606: 761-768. doi:10.1038/s41586-022-04835-6.

20. Shen B, Yi X, Sun Y, Bi X, Du J, Zhang C, et al. Proteomic and Metabolomic Characterization of COVID-19 Patient Sera. Cell. 2020; 182 (1): 59-72.e15. doi: 10.1016/j.cell.2020.05.032

21. Rössler T, Berezhnoy G, Singh Y, Cannet C, Reinsperger T, Schäfer H, Spraul M, Kneilling M, Merle U, Trautwein C. Quantitative Serum NMR Spectroscopy Stratifies COVID-19 Patients and Sheds Light on Interfaces of Host Metabolism and the Immune Response with Cytokines and Clinical Parameters. Metabolites. 2022; 12(12):1277. doi: 10.3390/metabo12121277

22. Ambikan AT, Yang H, Krishnan S, Svensson AS, Gupta S, Lourda M, et al. Multi-omics personalized network analyses highlight progressive disruption of central metabolism associated with COVID-19 severity. Cell Syst. 2022;13(8):665-681.e4. doi: 10.1016/j.cels.2022.06.006.

23. Lionetto L, Ulivieri M, Capi M, De Bernardini D, Fazio F, Petrucca, A;, et al. Increased kynurenine-to-tryptophan ratio in the serum of patients infected with SARS-CoV2: An observational cohort study. Biochim Biophys Acta Mol Basis Dis. 2021; 1867(3): 166042. doi: 10.1016/j.bbadis.2020.166042

24. Occelli C, Guigonis JM, Lindenthal S, Cagnard A, Graslin F, Brglez V, et al. Untargeted plasma metabolomic fingerprinting highlights several biomarkers for the diagnosis and prognosis of coronavirus disease 19. Front Med (Lausanne). 2022; 29(9):995069. doi: 10.3389/fmed.2022.995069.

25. Herrera Oostdam AS, Castañeda-Delgado JE, Oropeza-Valdez JJ, Borrego JC, Monárrez-Espino J, Zheng J, et al. Immunometabolic signatures predict risk of progression to sepsis in COVID-19. PLoS One. 2021;16(8):e0256784. doi: 10.1371/journal.pone.0256784.

26. Thomas T, Stefanoni D, Reisz JA, Nemkov T, Bertolone L, Francis RO, et al. COVID-19 infection alters kynurenine and fatty acid metabolism, correlating with IL-6 levels and renal status. JCI Insight. 2020; 23, 5(14):e140327. doi: 10.1172/jci.insight.140327.

27. Zhang Y, Guo R, Kim SH, Shah H, Zhang S, Liang JH, et al. SARS-CoV-2 hijacks folate and one-carbon metabolism for viral replication. Nat Commun. 2021; 12, 1676. doi: doi.org/10.1038/s41467-021-21903-z

28. Hwang TL, Shaka AJ. Water suppression that works. Excitation sculpting using arbitrary wave-forms and pulsed-field gradients. J Magn Reson. 1995; 112 (2): 275-279. doi: 10.1006/jmra.1995.1047

29. Carr HY, Purcell EM. Effects of Diffusion on Free Precession in Nuclear Magnetic Resonance Experiments. Phys Rev. 1954; 94: 630–638. doi: 10.1103/PhysRev.94.630

30. Ludwig C, Günther UL. MetaboLab - advanced NMR data processing and analysis for metabolomics. BMC Bioinformatics. 2011; 12, 366. doi:10.1186/1471-2105-12-366

31. Parsons HM, Ludwig C, Günther UL, Viant MR. Improved classification accuracy in 1- and 2-dimensional NMR metabolomics data using the variance stabilising generalised logarithm transformation. BMC Bioinformatics. 2007; 8, 234. doi: 10.1186/1471-2105-8-234

32. Bingol K, Li DW, Bruschweiler-Li L, Cabrera AO, Megraw T, Zhang F, et al. Unified and Isomer-Specific NMR Metabolomics Database for the Accurate Analysis of 13 C– 1 H HSQC Spectra. ACS Chem Biol. 2015; 10 (2):452–459. doi: 10.1021/cb5006382

33. Robinette SL, Zhang F, Brüschweiler-Li L, Brüschweiler R. Web Server Based Complex Mixture Analysis by NMR. Anal Chem. 2008; 80 (10):3606–3611. doi: 10.1021/ac702530t

34. Ulrich EL, Akutsu H, Doreleijers JF, Harano Y, Ioannidis YE, Lin J, et al. BioMagResBank. Nucleic Acids Res. 2007; 36 (1):D402–408. doi: 10.1093/nar/gkm957

35. Wishart, D.S.; Feunang, Y.D.; Marcu, A.; Guo, A.C.; Liang, K.; Vázquez-Fresno, R. et al. HMDB 4.0: the human metabolome database for 2018. Nucleic Acids Res. 2018; 46 (D1): D608– D617.

36. Kirwan JA, Gika H, Beger RD, Bearden D, Dunn WB, Goodacre R, et al. Quality assurance and quality control reporting in untargeted metabolic phenotyping: mQACC recommendations for analytical quality management. Metabolomics. 2022;18, 70. doi: 10.1007/s11306-022-01926-3.

37. Tsugawa H, Cajka T, Kind T, Ma Y, Higgins B, Ikeda K, et al. MS-DIAL: data-independent MS/MS deconvolution for comprehensive metabolome analysis. Nat. Methods. 2015; 12: 523–526. doi: 10.1038/nmeth.3393

38. Dunn WB, Broadhurst D, Begley P, et al. Procedures for large-scale metabolic profiling of serum and plasma using gas chromatography and liquid chromatography coupled to mass spectrometry. Nat. Protoc. 2011; 6 (7):1060–1083. doi: 10.1038/nprot.2011.335.

39. Tsugawa H, Kind T, Nakabayashi R, Yukihira D, Tanaka W, Cajka T, et al. Hydrogen Rearrangement Rules: Computational MS/MS Fragmentation and Structure Elucidation Using MS-FINDER Software. Anal. Chem. 2016; 88 (16): 7946–7958.

40. Broadhurst D, Goodacre R, Reinke SN, Kuligowski J, Wilson ID, Lewis MR, Dunn WB. Guidelines and considerations for the use of system suitability and quality control samples in mass spectrometry assays applied in untargeted clinical metabolomic studies. Metabolomics. 2018;14(6):72. doi: 10.1007/s11306-018-1367-3.

41. Chong J, Wishart DS, Xia J. Using MetaboAnalyst 4.0 for Comprehensive and Integrative Metabolomics Data Analysis. Cur Protoc Bioinformatics. 2019; 68(1):e86.doi: 10.1002/cpbi.86.

42. Breiman L, Friedman JH, Olshen RA, Stone CJ. Classification and Regression Trees; Routledge: New York, NY, 2017. 368 p.

43. Therneau T, Atkinson B, port BR (producer of the initial R, maintainer 1999-2017). rpart: Recursive Partitioning and Regression Trees 2022.

44. Ament Z, Bevers MB, Wolcott Z, Kimberly WT, Acharjee A. Uric Acid and Gluconic Acid as Predictors of Hyperglycemia and Cytotoxic Injury after Stroke. Transl Stroke Res. 2021; 12 (2): 293–302. doi: 10.1007/s12975-020-00862-5

45. Li T, Ning N, Li B, Luo D, Qin E, Yu W, Wang J, Yang G, Nan N, He Z, Yang N, Gong S, Li J, Liu A, Sun Y, Li Z, Jia T, Gao J, Zhang W, Huang Y, Hou J, Xue Y, Li D, Wei Z, Zhang L, Li B, Wang H. Longitudinal Metabolomics Reveals Ornithine Cycle Dysregulation Correlates With Inflammation and Coagulation in COVID-19 Severe Patients. Front Microbiol. 2021 Dec 3;12:723818. doi: 10.3389/fmicb.2021.723818.

46. Wen D, Zheng Z, Surapaneni A, Yu B, Zhou L, Zhou W, et al. Metabolite profiling of CKD progression in the chronic renal insufficiency cohort study. JCI Insight. 2022; 24,7(20):e161696. doi: 10.1172/jci.insight.161696.

47. Correia BSB, Ferreira VG, Piagge PMFD, Almeida MB, Assunção NA, Raimundo JRS, et al. ^1^H qNMR-Based Metabolomics Discrimination of Covid-19 Severity. J Proteome Res. 2022; 21, (7):1640-1653. https://doi.org/10.1021/acs.jproteome.1c00977

48. Kazak L, Rahbani JF, Samborska B, Lu GZ, Jedrychowski MP, Lajoie M, et al. Ablation of adipocyte creatine transport impairs thermogenesis and causes diet-induced obesity. Nat Metab. 2019, 1, 360-370. doi: 10.1021/acs.jproteome.1c00977

49. Kazak L, Cohen P. Creatine metabolism: energy homeostasis, immunity and cancer biology. Nat Rev Endocrinol. 2020; 16(8):421-436. doi: 10.1038/s41574-020-0365-5

50. Hu, S.; He, W.; Wu, G. Hydroxyproline in animal metabolism, nutrition, and cell signalling. Amino Acids. 2022; 54(4): 513–528. doi: 10.1007/s00726-021-03056-x

51. Ducker GS, Rabinowitz JD. One-Carbon Metabolism in Health and Disease. Cell Metab. 2017; 25(1): 27–42. doi: 10.1016/j.cmet.2016.08.009

52. Van der Veen JN, Kennelly JP, Wan S, Vance JE, Vance DE, Jacobs RL. The critical role of phosphatidylcholine and phosphatidylethanolamine metabolism in health and disease. Biochim Biophys Acta Biomembr. 2017, 1859, 1558–1572.

53. Silva RP, Eudy BJ, Deminice R. One-Carbon Metabolism in Fatty Liver Disease and Fibrosis: One-Carbon to Rule Them All. J Nutr. 2020; 150 (5): 994–1003. doi:10.1093/jn/nxaa032

54. Teixeira L, Temerozo JR, Pereira-Dutra FS, Ferreira AC, Mattos M, Gonçalves BS et al. Simvastatin Downregulates the SARS-CoV-2-Induced Inflammatory Response and Impairs Viral Infection Through Disruption of Lipid Rafts. Front Immunol. 2022, 13, 820131.

55. Lodge S, Nitschke P, Kimhofer T, Coudert JD, Begum S, Bong SH, et al. NMR Spectroscopic Windows on the Systemic Effects of SARS-CoV-2 Infection on Plasma Lipoproteins and Metabolites in Relation to Circulating Cytokines. J Proteome Res. 2021;20(2):1382-1396. doi: 10.1021/acs.jproteome.0c00876.

56. Wu D, Shu T, Yang X, Song JX, Zhang M, Yao C. et al. Plasma metabolomic and lipidomic alterations associated with COVID-19. Natl Sci Rev. 2020; 7(7): 1157–1168. doi: 10.1093/nsr/nwaa086

57. Roberts AB, Gu X, Buffa JA, Hurd AG, Wang Z, Zhu, W. et al. Development of a gut microbe-targeted nonlethal therapeutic to inhibit thrombosis potential. Nat Med. 2018; 24(9):1407–1417. doi: 10.1038/s41591-018-0128-1

58. Yang S, Li X, Yang F, Zhao R, Pan X, Liang J, et al. Gut Microbiota-Dependent Marker TMAO in Promoting Cardiovascular Disease: Inflammation Mechanism, Clinical Prognostic, and Potential as a Therapeutic Target. Front Pharmacol. 2019; 10:1360. doi: 10.3389/fphar.2019.01360

59. Manolis AS, Manolis TA, Manolis AA, Papatheou D, Melita H. COVID-19 Infection: Viral Macro- and Micro-Vascular Coagulopathy and Thromboembolism/Prophylactic and Therapeutic Management. J Cardiovasc Pharmacol Ther. 2021; 26(1): 12–24. doi: 10.1177/1074248420958973

60. Bennett JA, Mastrangelo MA, Ture SK, Smith CO, Loelius SG, Berg RA, et al. The choline transporter Slc44a2 controls platelet activation and thrombosis by regulating mitochondrial function. Nat Commun. 2020; 11, 3479. doi: 10.1038/s41467-020-17254-w

61. Sibal L, Agarwal SC, Home PD, Boger RH. The Role of Asymmetric Dimethylarginine (ADMA) in Endothelial Dysfunction and Cardiovascular Disease. Curr Cardiol Rev. 2010; 6 (2): 82–90. doi: 10.2174/157340310791162659

62. Zhao WC, Li G, Huang CY, Jiang JL. Asymmetric dimethylarginine: An crucial regulator in tissue fibrosis. Eur J Pharmacol. 2019; 854, 54–61. doi: 10.1016/j.ejphar.2019.03.055.

63. Ren W, Rajendran R, Zhao Y, Tan B, Wu G, Bazer FW, et al. Amino Acids As Mediators of Metabolic Cross Talk between Host and Pathogen. Front Immunol. 2018; 9, 319. doi: 10.3389/fimmu.2018.00319

64. Li P, Yin YL, Li D, Kim SW, Wu G. Amino acids and immune function. Br J Nutr. 2007, 98(2): 237–252. doi: 10.1017/S000711450769936X

65. Thiele I, Fleming RMT. Whole-body metabolic modelling predicts isoleucine dependency of SARS-CoV-2 replication. Comput Struct Biotechnol J. 2022; 20: 4098–4109. doi: 10.1016/j.csbj.2022.07.019

66. Oliveira LG, de Souza YA, Yamamoto P, Carregari VC, Crunfli F, Reis-de-Oliveira G, et al. SARS-CoV-2 infection impacts carbon metabolism and depends on glutamine for replication in Syrian hamster astrocytes. J Neurochem. 2022; 163(2): 113-132.

67. Páez-Franco JC, Maravillas-Montero JL, Mejía-Domínguez NR, Torres-Ruiz J, Tamez-Torres KM, Pérez-Fragoso A. Metabolomics analysis identifies glutamic acid and cystine imbalances in COVID-19 patients without comorbid conditions. Implications on redox homeostasis and COVID-19 pathophysiology. PLoS One. 2022; 17(9):e0274910. doi: 10.1371/journal.pone.0274910.

68. Frampas CF, Longman K, Spick M, Lewis HM, Costa CDS, Stewart A, et al. Untargeted saliva metabolomics by liquid chromatography-Mass spectrometry reveals markers of COVID-19 severity. PLoS One. 2022;17(9):e0274967. doi: 10.1371/journal.pone.0274967.

69. Bell JD, Brown JC, Nicholson JK, Sadler PJ. Assignment of resonances for “acute-phase” glycoproteins in high resolution proton NMR spectra of human blood plasma. FEBS Lett. 1987; 215(2): 311–315. doi: 10.1016/0014-5793(87)80168-0

70. Holmes E, Nicholson JK, Lodge S, Nitschke P, Kimhofer T, Wist J. Diffusion and relaxation edited proton NMR spectroscopy of plasma reveals a high-fidelity supramolecular biomarker signature of SARS-CoV-2 infection. Analytical Chemistry. 2021; 93(8): 3976–3986. https://doi.org/10.1021/acs.analchem.0c04952.

71. Lipman D, Safo SE, Chekouo T. Multi-omic analysis reveals enriched pathways associated with COVID-19 and COVID-19 severity. PLoS One. 2022;17(4):e0267047. doi: 10.1371/journal.pone.0267047.

72. Wang Y, Liu S, Liu H, Li W, Lin F, Jiang L, et al. SARS-CoV-2 infection of the liver directly contributes to hepatic impairment in patients with COVID-19. J Hepatol. 2020; 73 (4): 807–816. doi: 10.1016/j.jhep.2020.05.002

73. Lagana SM, Kudose S, Iuga AC, Lee MJ, Fazlollahi L, Remotti HE, et al. Hepatic pathology in patients dying of COVID-19: a series of 40 cases including clinical, histologic, and virologic data. Mod Pathol. 2020; 33, 2147–2155. doi: 10.1038/s41379-020-00649-x

74. Roberts I, Wright Muelas M, Taylor JM, Davison AS, Xu Y, Grixti JM, et al. Untargeted metabolomics of COVID-19 patient serum reveals potential prognostic markers of both severity and outcome. Metabolomics. 2021; 18, 6. doi: 10.1007/s11306-021-01859-3

75. Vázquez-Medina UM, Cerda-Reyes E, Galeana-Pavón A, López-Luna CE, Ramírez-Portillo PM, Ibañez-Cervantes G, et al. Interaction of metabolic dysfunction-associated fatty liver disease and nonalcoholic fatty liver disease with advanced fibrosis in the death and intubation of patients hospitalized with coronavirus disease 2019. Hepatol Commun. 2022; 6 (8): 2000–2010. doi: 10.1002/hep4.1957

76. Mooli RGR, Ramakrishnan SK. Emerging Role of Hepatic Ketogenesis in Fatty Liver Disease. Front Physiol. 2022; 13:946474. doi: 10.3389/fphys.2022.946474

77. Perazzo H, Cardoso SW, Ribeiro MPD, Moreira R, Coelho LE, Jalil EM, et al. In-hospital mortality and severe outcomes after hospital discharge due to COVID-19: A prospective multicenter study from Brazil. Lancet Reg Health Am. 2022; 11:100244. doi: 10.1016/j.lana.2022.100244. Erratum in: Lancet Reg Health Am. 2022.

78. Gebhard C, Regitz-Zagrosek V, Neuhauser HK, Morgan R, Klein SL. Impact of sex and gender on COVID-19 outcomes in Europe. Biol Sex Differ. 2020; 11, 29. doi: 10.1186/s13293-020-00304-9

79. Bechmann N, Barthel A, Schedl A, Herzig S, Varga Z, Gebhard C. et al. Sexual dimorphism in COVID-19: potential clinical and public health implications. Lancet Diabetes Endocrinol. 2022; 10 (3): 221–230. doi: 10.1016/S2213-8587(21)00346-6

